# Saliva is more sensitive for SARS-CoV-2 detection in COVID-19 patients than nasopharyngeal swabs

**DOI:** 10.1101/2020.04.16.20067835

**Authors:** Anne L. Wyllie, John Fournier, Arnau Casanovas-Massana, Melissa Campbell, Maria Tokuyama, Pavithra Vijayakumar, Bertie Geng, M. Catherine Muenker, Adam J. Moore, Chantal B.F. Vogels, Mary E. Petrone, Isabel M. Ott, Peiwen Lu, Arvind Venkataraman, Alice Lu-Culligan, Jonathan Klein, Rebecca Earnest, Michael Simonov, Rupak Datta, Ryan Handoko, Nida Naushad, Lorenzo R. Sewanan, Jordan Valdez, Elizabeth B. White, Sarah Lapidus, Chaney C. Kalinich, Xiaodong Jiang, Daniel J. Kim, Eriko Kudo, Melissa Linehan, Tianyang Mao, Miyu Moriyama, Ji Eun Oh, Annsea Park, Julio Silva, Eric Song, Takehiro Takahashi, Manabu Taura, Orr-El Weizman, Patrick Wong, Yexin Yang, Santos Bermejo, Camila Odio, Saad B. Omer, Charles S. Dela Cruz, Shelli Farhadian, Richard A. Martinello, Akiko Iwasaki, Nathan D. Grubaugh, Albert I. Ko

## Abstract

Rapid and accurate SARS-CoV-2 diagnostic testing is essential for controlling the ongoing COVID-19 pandemic. The current gold standard for COVID-19 diagnosis is real-time RT-PCR detection of SARS-CoV-2 from nasopharyngeal swabs. Low sensitivity, exposure risks to healthcare workers, and global shortages of swabs and personal protective equipment, however, necessitate the validation of new diagnostic approaches. Saliva is a promising candidate for SARS-CoV-2 diagnostics because (1) collection is minimally invasive and can reliably be self-administered and (2) saliva has exhibited comparable sensitivity to nasopharyngeal swabs in detection of other respiratory pathogens, including endemic human coronaviruses, in previous studies. To validate the use of saliva for SARS-CoV-2 detection, we tested nasopharyngeal and saliva samples from confirmed COVID-19 patients and self-collected samples from healthcare workers on COVID-19 wards. When we compared SARS-CoV-2 detection from patient-matched nasopharyngeal and saliva samples, we found that saliva yielded greater detection sensitivity and consistency throughout the course of infection. Furthermore, we report less variability in self-sample collection of saliva. Taken together, our findings demonstrate that saliva is a viable and more sensitive alternative to nasopharyngeal swabs and could enable at-home self-administered sample collection for accurate large-scale SARS-CoV-2 testing.

## Introduction

Efforts to control SARS-CoV-2, the novel coronavirus causing COVID-19 pandemic, depend on accurate and rapid diagnostic testing. These tests must be (***1***) sensitive to mild and asymptomatic infections to promote effective self isolation and reduce transmission within high risk groups^1^; (***2***) consistent to reliably monitor disease progression and aid clinical decisions^2^; and (***3***) scalable to inform local and national public health policies, such as when social distancing measures can be safely relaxed. However, current SARS-CoV-2 testing strategies often fail to meet these criteria, in part because of their reliance on nasopharyngeal swabs as the widely recommended sample type for real-time RT-PCR. Although nasopharyngeal swabs are commonly used in respiratory virus diagnostics, they show relatively poor sensitivity for SARS-CoV-2 detection in early infection and are inconsistent during serial testing^2–6^. Moreover, collecting nasopharyngeal swabs causes discomfort to patients due to the procedure’s invasiveness, limiting compliance for repeat testing, and presents a considerable risk to healthcare workers, because it can induce patients to sneeze or cough, expelling virus particles^7^. The procedure is also not conducive to large-scale testing, because there are widespread shortages of swabs and personal protective equipment for healthcare workers^8^, and self-collection of nasopharyngeal swabs is difficult and less sensitive for virus detection^9^. These challenges will be further exacerbated as the COVID-19 pandemic intensifies in low income countries. Given the limitations, a more reliable and less resource-intensive sample collection method, ideally one that accommodates self-collection in the home, is urgently needed.

Saliva sampling is an appealing alternative to nasopharyngeal swab, since collecting saliva is non-invasive and easy to self-administer. An analysis of nasopharyngeal and saliva concordance for RT-PCR detection of respiratory pathogens, including two seasonal human coronaviruses, suggests comparable diagnostic sensitivity between the two sample types^10,11^. Preliminary findings indicate that (***1***) SARS-CoV-2 can be detected from the saliva of COVID-19 patients^12^ and (***2***) self-collected saliva samples have comparable SARS-CoV-2 detection sensitivity to nasopharyngeal swabs collected by healthcare workers from mild and subclinical COVID-19 cases^13^. Critically, however, no rigorous evaluation of the sensitivity of SARS-CoV-2 detection in saliva with respect to nasopharyngeal swabs has been conducted from inpatients during the course of COVID-19 infection.

In this study, we evaluated SARS-CoV-2 detection in paired nasopharyngeal swabs and saliva samples collected from COVID-19 inpatients and asymptomatic healthcare workers at moderate-to-high risk of COVID-19 exposure. Our results indicate that using saliva for SARS-CoV-2 detection is more sensitive and consistent than using nasopharyngeal swabs. Overall, we demonstrate that saliva should be considered as a reliable sample type to alleviate COVID-19 testing demands.

## Results

### Higher SARS-CoV-2 titers detected from saliva than nasopharyngeal swabs from inpatients

To determine if saliva performs as well as the U.S. CDC recommendation of using nasopharyngeal swabs for SARS-CoV-2 diagnostics, we collected clinical samples from 44 COVID-19 inpatient study participants (**Table 1**). This cohort represents a range of COVID-19 patients with severe disease, with 19 (43%) requiring intensive care, 10 (23%) requiring mechanical ventilation, and 2 (5%) deceased as of April 5th, 2020. Using the U.S. CDC SARS-CoV-2 RT-PCR assay, we tested 121 self-collected saliva or healthcare worker-administered nasopharyngeal swabs from this cohort. We found strong concordance between the U.S. CDC “N1” and “N2” primer-probe sets (**Extended Data Fig. 1**), and thus calculated virus titers (virus copies/mL) using only the “N1” set. From all positive samples tested (*n* = 46 nasopharyngeal, 37 saliva), we found that the geometric mean virus titers from saliva were about 5X higher than nasopharyngeal swabs (*p* < 0.05, Mann-Whitney test; **Fig. 1a**). When limiting our analysis to only patient-matched nasopharyngeal and saliva samples (*n* = 38 for each sample type), we found that SARS-CoV-2 titers from saliva were significantly higher than nasopharyngeal swabs (*p* = 0.0001, Wilcoxon test; **Fig. 1b**). Moreover, we detected SARS-CoV-2 from the saliva but not the nasopharyngeal swabs from eight matching samples (21%), while we only detected SARS-CoV-2 from nasopharyngeal swabs and not saliva from three matched samples (8%; **Fig. 1c**). Overall, we found higher SARS-CoV-2 titers from saliva than nasopharyngeal swabs from hospital inpatients.

**Table 1.**
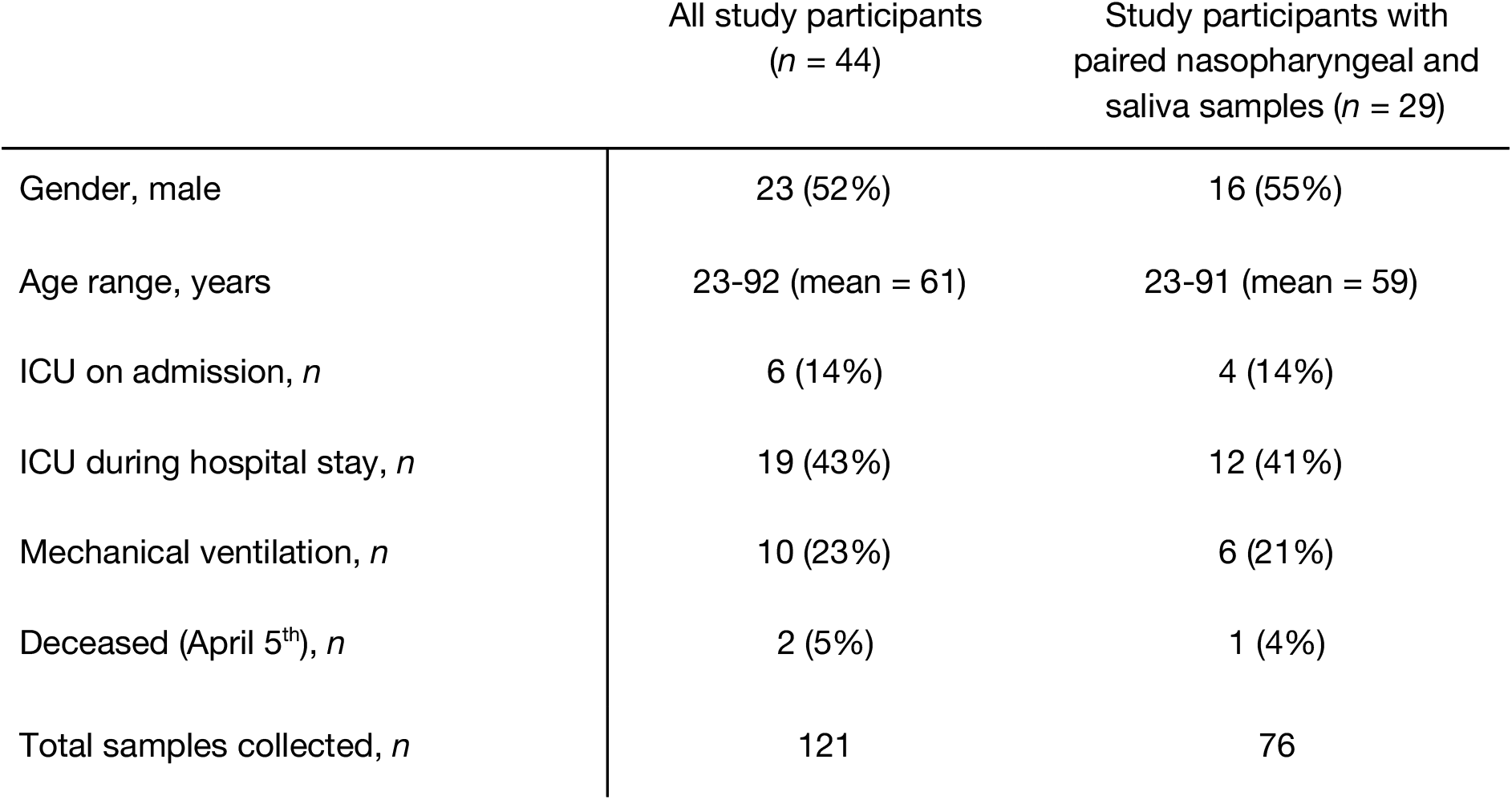
COVID-19 inpatient cohort characteristics

| | All study participants<br>($n = 44$) | Study participants with<br>paired nasopharyngeal and<br>saliva samples ($n = 29$) |
| --- | --- | --- |
| Gender, male | 23 (52%) | 16 (55%) |
| Age range, years | 23-92 (mean = 61) | 23-91 (mean = 59) |
| ICU on admission, $n$ | 6 (14%) | 4 (14%) |
| ICU during hospital stay, $n$ | 19 (43%) | 12 (41%) |
| Mechanical ventilation, $n$ | 10 (23%) | 6 (21%) |
| Deceased (April 5<sup>th</sup>), $n$ | 2 (5%) | 1 (4%) |
| Total samples collected, $n$ | 121 | 76 |

**Figure 1.**
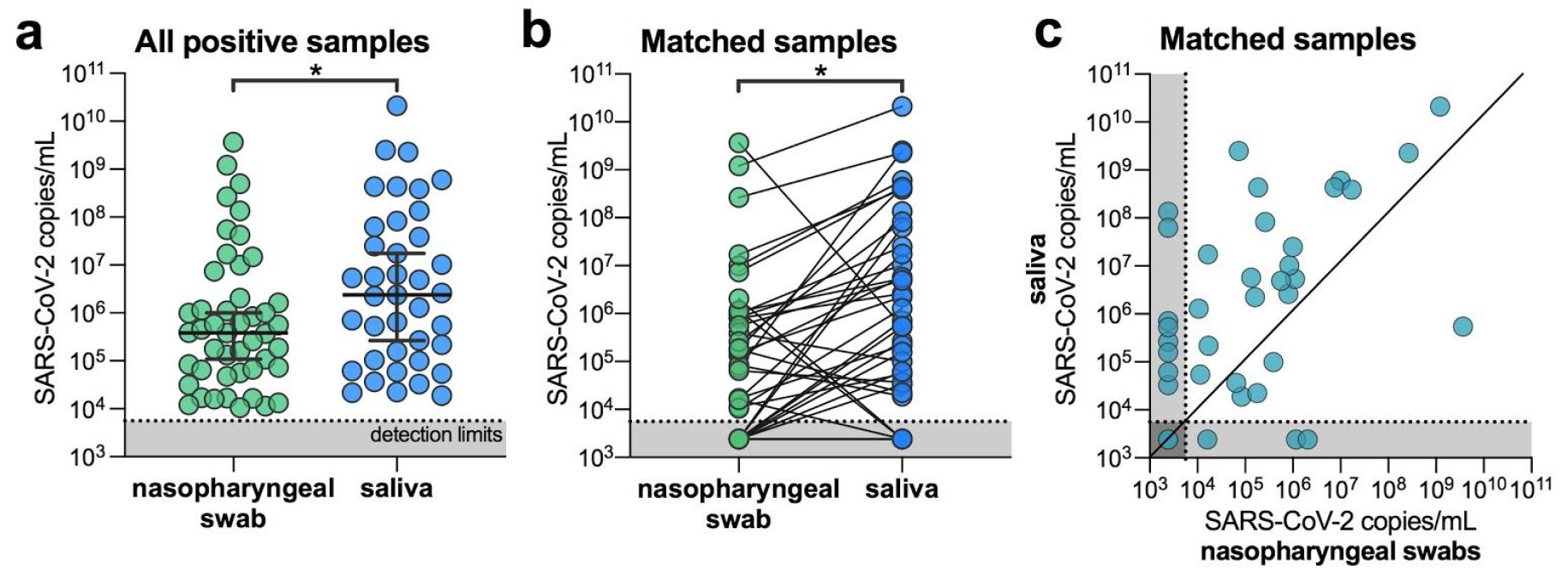
SARS-CoV-2 titers are higher in the saliva than nasopharyngeal swabs from hospital inpatients. (**a**) All positive nasopharyngeal swabs (*n* = 46) and saliva samples (*n* = 39) were compared by a Mann-Whitney test (*p* < 0.05). Bars represent the median and 95% CI. Our assay detection limits for SARS-CoV-2 using the US CDC “N1” assay is at cycle threshold 38, which corresponds to 5,610 virus copies/mL of sample (shown as dotted line and grey area). (**b**) Patient matched samples (*n* = 38), represented by the connecting lines, were compared by a Wilcoxon test test (*p* < 0.05). (**c**) Patient matched samples (*n* = 38) are also represented on a scatter plot. All of the data used to generate this figure, including the raw cycle thresholds, can be found in **Supplementary Data 1. Extended Data Fig. 1** shows the correlation between US CDC assay “N1” and “N2” results.

### Less temporal SARS-CoV-2 variability when testing saliva from inpatients

As temporal SARS-CoV-2 diagnostic testing from nasopharyngeal swabs is reported to be variable^2,3^, we tested longitudinal nasopharyngeal and saliva samples from inpatients to determine which sample type provided more consistent results. From 22 participants with multiple nasopharyngeal swabs and 12 participants with multiple saliva samples, we found that SARS-CoV-2 titers generally decreased in both sample types following the reported date of symptom onset (**Fig. 2a**). Our nasopharyngeal swab results are consistent with previous reports of variable SARS-CoV-2 titers and results^2,3^: we found 5 instances where a participant’s nasopharyngeal swab was negative for SARS-CoV-2 followed by a positive result during the next collection (5/33 repeats, 33%; **Fig. 2b**). In longitudinal saliva collections from 12 patients, however, there were no instances in which a sample tested negative and was later followed by a positive result. As true negative test results are important for clinicians to track patient improvements and for decisions regarding discharges, our data suggests that saliva is a more consistent sample type than nasopharyngeal swabs for monitoring temporal changes in SARS-CoV-2 titers.

**Figure 2:**
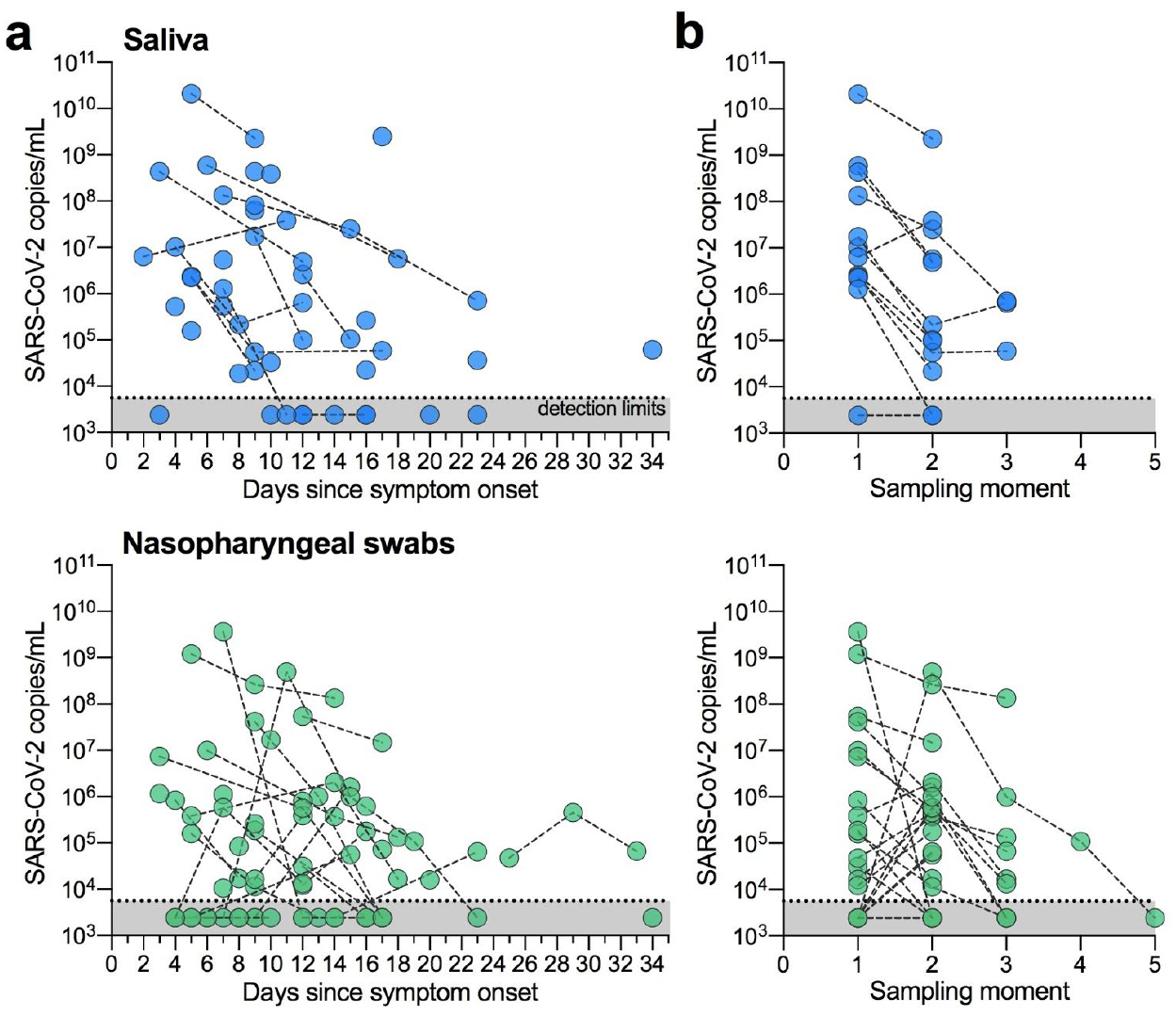
SARS-CoV-2 detection is less variable between repeat sample collections with saliva. (a) Longitudinal SARS-CoV-2 titers from saliva or nasopharyngeal swabs are shown as days since symptom onset. Each circle represents a separate sample, which are connected to additional samples from the same patient by a dashed line. Our assay detection limits for SARS-CoV-2 using the US CDC “N1” assay is at cycle threshold 38, which corresponds to 5,610 virus copies/mL of sample (shown as dotted line and grey area). (**b**) The data are also shown by sampling moment (sequential collection time) to highlight the differences in virus titers between collection points. All of the data used to generate this figure, including the raw cycle thresholds, can be found in **Supplementary Data 1**.

### More consistent self-sampling from healthcare workers using saliva

Validating saliva for the detection of subclinical SARS-CoV-2 infections could prove transformative for both remote patient diagnostics and healthcare worker surveillance. To investigate this, we enrolled 98 asymptomatic healthcare workers into our study and collected saliva and/or nasopharyngeal swabs on average every 2.9 days (range = 1-8 days, **Table 2**). To date, we have detected SARS-CoV-2 in saliva from two healthcare workers who were negative by nasopharyngeal swabs using both the US CDC “N1” and “N2” tests and did not report any symptoms. The saliva from one of these individuals again tested positive alongside a matching negative nasopharyngeal swab upon repeat testing 2 days later. Virus titers from asymptomatic healthcare workers’ saliva are lower than what we typically detect from symptomatic inpatients (**Fig. 3a**), which likely supports the lack of symptoms.

**Table 2.** Healthcare worker cohort

|  | All study participants<br>(n=98) | Participants with<br>matching samples (n=33) |
| --- | --- | --- |
| Gender, male | 16 (16%) | 5 (15%) |
| Age range, years (average) | 22-67 (36) | 22-61 (36) |
| Total samples collected, n | 244 | 64 |

**Figure 3.**
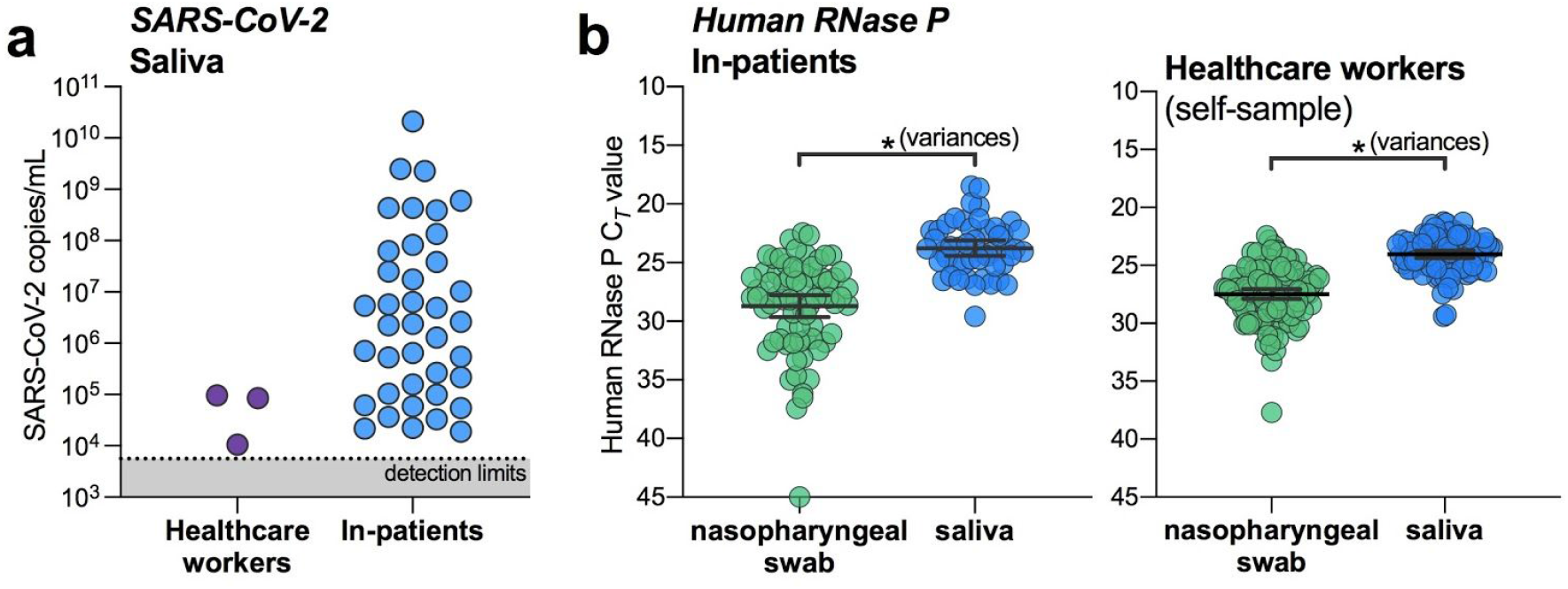
Saliva is an alternative for SARS-CoV-2 screening from healthcare workers and asymptomatic cases. (**a**) SARS-CoV-2 titers measured from the saliva of healthcare workers and inpatients. Our assay detection limits for SARS-CoV-2 using the US CDC “N1” assay is at cycle threshold 38, which corresponds to 5,610 virus copies/mL of sample (shown as dotted line and grey area). (**b**) RT-PCR cycle thresholds (Ct) values for human RNase P, and internal control for sample collection, from either inpatients (left panel) or health care workers (right panel) were compared by variances using the F test (*p* = 0.0001 for inpatients; *p* = 0.0002 for healthcare workers). All of the data used to generate this figure, including the raw cycle thresholds, can be found in **Supplementary Data 1. Table 1. COVID-19 inpatient cohort characteristics**

Our limited data supports that saliva may be more sensitive for detecting asymptomatic or pre-symptomatic infections; however, a larger sample size is needed to confirm. As nasopharyngeal swab sampling inconsistency may be one of the potential issues for false negatives (**Fig. 2**), monitoring an internal control for proper sample collection, human RNase P, may provide an alternative evaluation technique. While human RNase P detection was better from saliva in both the inpatient and healthcare worker cohorts (**Fig. 3b**), this alone may not indicate better virus detection. More importantly, we found that human RNase P detection was more variable from nasopharyngeal swabs collected from inpatients (*p* = 0.0001, F test for variances) and self-collected from healthcare workers (*p* = 0.0002; **Fig. 3b**). Our results suggest that saliva may also be an appropriate, and perhaps more sensitive, alternative to nasopharyngeal swabs for screening asymptomatic or pre-symptomatic SARS-CoV-2 infections.

## Discussion

Our study demonstrates that saliva is a viable and preferable alternative to nasopharyngeal swabs for SARS-CoV-2 detection. We found that the sensitivity of SARS-CoV-2 detection from saliva is comparable, if not superior to nasopharyngeal swabs in early hospitalization and is more consistent during extended hospitalization and recovery. Moreover, the detection of SARS-CoV-2 from the saliva of two asymptomatic healthcare workers despite negative matched nasopharyngeal swabs suggests that saliva may also be a viable alternative for identifying mild or subclinical infections. With further validation, widespread implementation of saliva sampling could be transformative for public health efforts: saliva self-collection negates the need for direct healthcare worker-patient interaction, a source of several major testing bottlenecks and overall nosocomial infection risk^14–16^, and alleviates supply demands on swabs and personal protective equipment.

As SARS-CoV-2 viral loads differ between mild and severe cases^17^, a limitation of our study is the primary focus on COVID-19 inpatients, many with severe disease. While more data are required to more rigorously compare the efficacy of saliva in the hospital setting to earlier in the course of infection, findings from two recent studies support its potential for detecting SARS-CoV-2 from both asymptomatic individuals and outpatients^13,18^. As infectious virus has been detected from the saliva of COVID-19 patients^12^, ascertaining the relationship between virus genome copies and infectious virus particles in the saliva of pre-symptomatic individuals^19^ will play a key role in understanding the dynamics of asymptomatic transmission^1,20^.

Stemming from the promising results for SARS-CoV-2 detection in asymptomatic individuals^13^, a saliva SARS-CoV-2 detection assay has already gained approval through the U.S. Food and Drug Administration emergency use authorization^18^. To meet the growing testing demands, however, our findings support the need for immediate validation and implementation of saliva for SARS-CoV-2 diagnostics in certified clinical laboratories.

## Methods

### Ethics

All study participants were enrolled and sampled in accordance to the Yale University HIC-approved protocol #2000027690. Demographics, clinical data and samples were only collected after the study participant had acknowledged that they had understood the study protocol and signed the informed consent. All participant information and samples were collected in association with study identifiers.

### Participant enrollment

#### Inpatients

Patients admitted to Yale New Haven Hospital (a 1541-bed tertiary care medical center in New Haven, CT, USA), who tested positive for SARS-CoV-2 by nasopharyngeal and/or oropharyngeal swab (CDC approved assay) were invited to enroll in the research study. Exclusion criteria were age under 18 years, non-English speaking and clinical, radiological or laboratory evidence for a non-infectious cause of fever or respiratory symptoms or a microbiologically-confirmed infectious source (e.g. gastrointestinal, urinary, cardiovascular) other than respiratory tract for symptoms and no suspicion for COVID-19 infection.

#### Healthcare workers

Asymptomatic healthcare workers (e.g., without fever or respiratory symptoms) with occupational exposure to patients with COVID-19 were invited to enroll in the study. Study participation enabled active surveillance to ensure early detection following exposure and to further protect other healthcare workers and patients.

## Sample collection

### Inpatients

Nasopharyngeal and saliva samples were obtained every three days throughout their clinical course. Nasopharyngeal samples were taken by registered nurses using the BD universal viral transport (UVT) system. The flexible, mini-tip swab was passed through the patient’s nostril until the posterior nasopharynx was reached, left in place for several seconds to absorb secretions then slowly removed while rotating. The swab was placed in the sterile viral transport media (total volume 3 mL) and sealed securely. Saliva samples were self-collected by the patient. Upon waking, patients were asked to avoid food, water and brushing of teeth until the sample was collected. Patients were asked to repeatedly spit into a sterile urine cup until roughly a third full of liquid (excluding bubbles), before securely closing it. All samples were stored at room temperature and transported to the research lab at the Yale School of Public Health within 5 hours of sample collection.

### Healthcare workers

Healthcare workers were asked to collect a self-administered nasopharyngeal swab and a saliva sample every three days for a period of 2 weeks. Samples were stored at +4°C until being transported to the research lab.

## SARS-CoV-2 detection

On arrival at the research lab, total nucleic acid was extracted from 300 µl of viral transport media from the nasopharyngeal swab or 300 µl of whole saliva using the MagMAX Viral/Pathogen Nucleic Acid Isolation kit (ThermoFisher Scientific) following the manufacturer’s protocol and eluted into 75 µl of elution buffer. For SARS-CoV-2 RNA detection, 5 µl of RNA template was tested as previously described^21,22^, using the US CDC real-time RT-PCR primer/probe sets for 2019-nCoV_N1 and 2019-nCoV_N2 and the human RNase P (RP) as an extraction control. Samples were classified as positive for SARS-CoV-2 when both N1 and N2 primer-probe sets were detected <38 C_*T*_. Virus copies were quantified using a 10-fold dilution standard curve of RNA transcripts that we previously generated^21^. As results from N1 and N2 were comparable (**Extended Data Fig. 1**), all virus copies are shown as calculated using the N1 primer-probe set.

## Statistical analysis

Statistical analyses were conducted in GraphPad Prism 8.0.0 as described in the Results.

## Data Availability

All data generated and/or analysed during the current study are available from the corresponding author on reasonable request.

## Acknowledgments

We gratefully acknowledge the study participants for their time and commitment to the study. We thank all members of the clinical team at Yale-New Haven Hospital for their dedication and work which made this study possible. We also thank S. Taylor and P. Jack for technical discussions.

## Extended data

**Extended Data Fig. 1.**
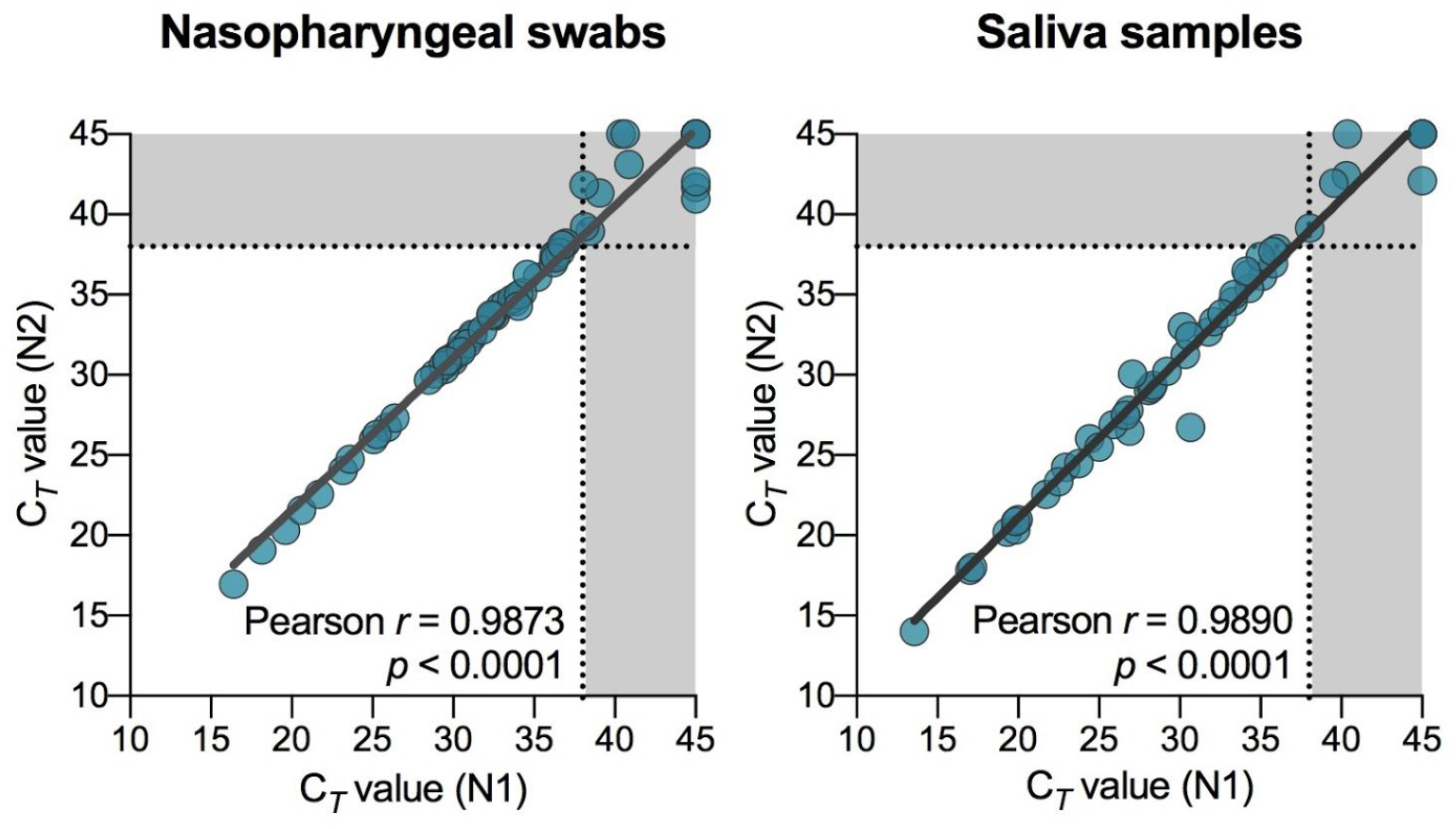
Concordance between SARS-CoV-2 detection using US CDC “N1” and “N2” primer and probe sets. Ct = RT-PCR cycle threshold. Dotted line and grey areas indicate the limits of detection.

